# Prediction of Maternal Hemorrhage: Using Machine Learning to Identify Patients at Risk

**DOI:** 10.1101/2020.06.04.20122663

**Authors:** Jill M. Westcott, Francine Hughes, Wenke Liu, Mark Grivainis, David Fenyö

## Abstract

**Background:** Postpartum hemorrhage remains one of the largest causes of maternal morbidity and mortality in the United States.

**Objective:** To utilize machine learning techniques to identify patients at risk for postpartum hemorrhage at obstetric delivery.

**Study Design:** Women aged 18 to 55 delivering at a major academic center from July 2013 to October 2018 were included for analysis (n = 30,867). A total of 497 variables were collected from the electronic medical record including demographic information, obstetric, medical, surgical, and family history, vital signs, laboratory results, labor medication exposures, and delivery outcomes. Postpartum hemorrhage was defined as a blood loss of ≥ 1000 mL at the time of delivery, regardless of delivery method, with 2179 positive cases observed (7.06%).

Supervised learning with regression-, tree-, and kernel-based machine learning methods was used to create classification models based upon training (n = 21,606) and validation (n = 4,630) cohorts. Models were tuned using feature selection algorithms and domain knowledge. An independent test cohort (n = 4,631) determined final performance by assessing for accuracy, area under the receiver operating curve (AUC), and sensitivity for proper classification of postpartum hemorrhage. Separate models were created using all collected data versus limited to data available prior to the second stage of labor/at the time of decision to proceed with cesarean delivery. Additional models examined patients by mode of delivery.

**Results:** Gradient boosted decision trees achieved the best discrimination in the overall model. The model including all data mildly outperformed the second stage model (AUC 0.979, 95% CI 0.971–0.986 vs. AUC 0.955, 95% CI 0.939–0.970). Optimal model accuracy was 98.1% with a sensitivity of 0.763 for positive prediction of postpartum hemorrhage. The second stage model achieved an accuracy of 98.0% with a sensitivity of 0.737. Other selected algorithms returned ≥ models that performed with decreased discrimination. Models stratified by mode of delivery achieved good to excellent discrimination, but lacked sensitivity necessary for clinical applicability.

**Conclusions:** Machine learning methods can be used to identify women at risk for postpartum hemorrhage who may benefit from individualized preventative measures. Models limited to data available prior to delivery perform nearly as well as those with more complete datasets, supporting their potential utility in the clinical setting. Further work is necessary to create successful models based upon mode of delivery. An unbiased approach to hemorrhage risk prediction may be superior to human risk assessment and represents an area for future research.

**Condensation:** Machine learning methods can be successfully utilized to predict nearly three-quarters of women at risk of postpartum hemorrhage when undergoing obstetric delivery.

**AJOG at a Glance:**

A. Why was the study conducted?

- To determine patients at risk for postpartum hemorrhage using modern machine learning techniques on a robust data set directly derived from the electronic medical record
B. What are the key findings?

- Using 28 predictor features, the model successfully classified 73.7% of patients who ultimately had a postpartum hemorrhage using information available prior to delivery
- Many previously identified risk factors for postpartum hemorrhage were not included in the final model, potentially discounting their contribution to hemorrhage risk
- Models stratified by delivery method achieved good to excellent discrimination but noted lower sensitivity and need further investigation
C. What does this study add to what is already known?

- This study represents the largest cohort directly-derived from the electronic medical record to use machine learning techniques to identify patients at risk for postpartum hemorrhage

## Introduction

Postpartum hemorrhage is the leading cause of maternal mortality worldwide^1^. In the United States, the rate of postpartum hemorrhage continues to rise, complicating nearly 3% of deliveries^2^. Mothers with severe hemorrhage may require blood transfusion, hysterectomy, or intensive care unit admission with a select number of cases proving fatal. Postpartum hemorrhage that leads to blood transfusion is the leading cause of severe maternal morbidity in the United States^3^. Stewardship of blood resources and minimizing hemorrhage related morbidity remain ongoing efforts as blood transfusion is not without risk. By predicting patients at risk for significant blood loss, prophylactic measures may be instituted to avoid maternal morbidity and mortality.

A number of risk factors for postpartum hemorrhage have been established, including previous postpartum hemorrhage, multifetal gestation, pre-eclampsia, augmented labor, fetal macrosomia, operative vaginal delivery, and complex lacerations, as well as other factors^4^. Previous models for prediction of postpartum hemorrhage have been developed^5,6,7^, but validation of these among different populations and at different time points within the labor process has been limited. A machine learning study using administrative data provided poor discrimination for predicting need for hospital readmission due to postpartum in the first 12 weeks postpartum^8^. Prediction of postpartum hemorrhage remains a challenge for the obstetric provider and further work is necessary using modern modeling methods.

The field of machine learning has recently seen a fast development of methods that support unbiased learning from data. Supervised learning involves processing information to predict from examples with a known outcome, often for the purpose of estimating risk in examples where the outcome is not known^9^. Multiple applications for machine learning exist within medicine, but to date have not been widely utilized in the field of obstetrics. By using the power of modern predictive modeling for postpartum hemorrhage, we aim to better identify those patients at increased risk for obstetric hemorrhage to avoid maternal morbidity and mortality. Identifying those patients at highest risk of postpartum hemorrhage will enable providers to reduce the cost and morbidity associated with postpartum hemorrhage and ultimately improve patient outcomes.

## Material and Methods

This was a retrospective cohort study conducted at a single tertiary care center. Women aged 18 to 55 delivering at New York University Langone Health Tisch Hospital from July 1, 2013, to October 31, 2018, were included for analysis. Patients not meeting age parameters as well as those cases in which a blood loss value was either not available or not recorded were excluded. Institutional Review Board approval was obtained from New York University Langone Health.

A total of 497 variables were collected from unique sources within the electronic medical record including demographic information, obstetric, medical, surgical, and family history, vital signs, laboratory results, labor exposures, and delivery outcomes. Postpartum hemorrhage was defined as a blood loss of ≥ 1000 mL at the time of delivery as recommended by the ACOG revitalize program^10^.

The delivery cohort was split into training (70%) and validation (15%) sets for model creation. Supervised learning with regression-, tree-, and kernel-based machine learning methods was used to create classification models. Models were tuned using recursive feature selection, selection by filtering, observing feature importance, and domain knowledge. Model parameters were customized and examined to produce optimal results. An independent test cohort (15%) determined final performance by assessing for accuracy, area under the receiver operating curve (AUC), and sensitivity for proper classification of postpartum hemorrhage.

The initial model included variables that contained information that would be feasible to obtain prior to delivery (i.e. relevant historical information, objective data present within the inpatient and outpatient chart, and diagnoses associated with the patient’s delivery encounter entered within 24 hours following delivery). A secondary model was created limited to data strictly expected to be available prior to the second stage of labor/at the time of decision to proceed with cesarean delivery, as this was likely the more clinically useful tool. Additional models were created for patients undergoing cesarean and vaginal delivery.

Selection of appropriate variables for inclusion was made by an obstetric provider with experience and knowledge of the electronic medical record. Variables were processed according to the provider’s assessment of the clinical scenario noted for each patient.

## Results

A total of 30,867 patients met inclusion criteria and 2179 (7.06%) cases met criteria for postpartum hemorrhage. The average gestational age was 274.6 days (range 107–303) and average patient age was 32.7 (range 18–55). Cesarean delivery was noted in 27.6% (n = 8,534) patients with the remainder undergoing vaginal delivery. The delivery cohort was split into training, validation, and test cohorts for initial model creation containing 21,606, 4,630, and 4,631 patients, respectively.

The initial model included a total of 280 variables. Logistic regression, Random Forest, gradient boosted decision trees (XGBoost), and Support Vector Machine models were generated to create a representative sample of different methods. Gradient boosted decision trees achieved the best discrimination among the initial models, performing with an AUC of 0.979 (95% CI 0.971–0.986) and an accuracy of 98.1%. Sensitivity for this model was 0.763 (Table 1). Other models performed less successfully. The optimal model included 212 features.

**Table 1:** Optimal performance of all models using gradient boosted decision trees

| **Model** | **Accuracy** | **AUC** | **Sensitivity** |
| --- | --- | --- | --- |
| Initial | 0.981 | 0.979 | 0.763 |
| Second stage | 0.980 | 0.955 | 0.737 |
| Cesarean delivery | 0.818 | 0.737 | 0.320 |
| Vaginal delivery (one hour) | 0.982 | 0.837 | 0.254 |
| Vaginal delivery (second stage) | 0.983 | 0.846 | 0.254 |

The dataset was then trimmed to include only those variables (123 in total) available prior to the second stage of labor or at the time of decision to proceed with cesarean delivery. A similar representative sample of modeling methods was used, with gradient boosted decision trees again achieving the best discrimination, noting an AUC of 0.955 (95% CI 0.939–0.970) and an accuracy of 98.0%. Sensitivity for this model was 0.737 (Table 1). This model included a total of 28 features (Table 2). The most important features included body mass index, admission hematocrit, cesarean delivery prior to labor/rupture, scheduling status of cesarean delivery, and admission platelet count.

**Table 2:** Features including in optimal second stage model

| Non-laboratory components | Laboratory components |
| --- | --- |
| Patient age | Eosinophils (absolute) |
| Gestational age | Hematocrit* |
| Systolic blood pressure | Hemoglobin |
| Diastolic blood pressure | Lymphocytes (absolute) |
| Body mass index* | Lymphocytes (percent) |
| Pulse oximetry | Mean corpuscular hemoglobin |
| Temperature | Mean corpuscular hemoglobin concentration |
| Live birth count | Mean corpuscular volume |
| Baseline fetal heart rate | Mean platelet volume |
| Amniotic fluid color | Monocytes (percent) |
| Cesarean delivery prior to labor/rupture* | Neutrophils (absolute) |
| Cesarean delivery scheduling status* | Neutrophils (percent) |
|  | Platelet count* |
|  | Red cell distribution width (standard deviation) |
|  | Red blood cell count |
|  | White blood cell count |

301 \*Top 5 in feature importance

Additional models focused on classifying patients by mode of delivery. A model examining those patients ultimately delivered by cesarean delivery (n = 8,534) was created using information available at the time of decision to proceed with cesarean delivery (i.e. within 1 hour after presentation for scheduled procedure or following attempted vaginal delivery prior to proceeding to the operating room). A total of 173 variables were included. Using gradient boosted decision trees, the highest performing model contained 76 variables and achieved an AUC of 0.737 (95% CI 0.703–0.772). Accuracy of 81.8% and a lower sensitivity of 0.320 were noted (Table 1).

Two models using those patients who underwent vaginal delivery (n = 22,333) were also created. The first focused on information available within 1 hour of admission for presumed vaginal delivery and examined 127 variables. The model achieved excellent discrimination, noting an AUC of 0.837 (95% CI 0.782–0.893) and accuracy of 98.2%; however, the sensitivity remained low at 0.254. The optimal model included 7 variables. The second model utilized information available prior to the second stage of labor, thus included variables related to the patient’s labor course, including medication exposure and induction method, if applicable. A total of 176 features were included, resulting in an optimal model with AUC 0.846 (95% CI 0.790–0.902) and accuracy 98.3%. Sensitivity was again 0.254 and the final model included 92 features (Table 1). Both of these models were achieved using gradient boosted decision trees. Third trimester and admission hemoglobin/hematocrit were among the most important features in both vaginal delivery models.

## Structured Discussion/Comment

### Principal Findings

Our study has successfully produced a model for predicting postpartum hemorrhage in patients undergoing vaginal delivery. When using only the data available prior to the second stage of labor or at the time to proceed with vaginal delivery, we achieved nearly equal discrimination and sensitivity compared to our more robust initial model, successfully predicting nearly 3 out of every 4 patients who had a postpartum hemorrhage.

Many previously identified risk factors for postpartum hemorrhage were not included in the final model, including multiple gestation, operative vaginal delivery, and history of postpartum hemorrhage, among others. This indicates that many of these factors may not be as contributory to postpartum hemorrhage risk as previously believed, but further work is necessary.

### Results

Postpartum hemorrhage is a known cause of significant maternal morbidity and mortality in the United States and remains difficult to predict. Few existing studies have utilized machine learning methods to identify patients at risk for postpartum hemorrhage with minimal success^5,6,7,8^. A recently published model used a large cohort from the U.S Consortium for Safe Labor and achieved excellent discrimination, although its utility in the clinical setting is limited^11^. This study used 55 predictor variables, indicating a less robust data set than curated for our model. Our study represents the largest cohort to date to generate a predictive risk model using data directly abstracted from the electronic medical record that is applicable in a targeted population.

When stratified by delivery method, our models noted a decreased sensitivity. While this may appear in contradiction to expected results, it is understandable due to the fact that the majority of postpartum hemorrhages occurred in those patients who underwent cesarean delivery. This is further reflected by examining the most important features in our final second stage model.

### Clinical Implications

The ability to predict patients at risk for postpartum hemorrhage using readily available information represents an area of tremendous clinical opportunity. Integrating a model such as ours into clinical practice will give providers the real-time capability to assess a patient’s risk of hemorrhage. Targeted intervention, such as prophylactic administration of uterotonic medication, availability of blood products, and even potentially transfer to a center offering a higher level of maternal care^12^ is a consideration for those patients deemed at risk.

### Research Implications

In machine learning, models can continuously be tuned with new data, enabling further potential improvement in predictive capability. The amount of data available due to the advent of the electronic medical record is vast, hence the use of these methods in obstetrics is in its infancy. Continued exploration of different modeling and data abstraction techniques will be key to using machine learning to benefit our field.

This specific study represents the first large-scale predictive model using machine learning for use in a targeted population with largely positive results. At our institution, discussions have begun surrounding implementation of this model as a part of routine obstetric management. Continued model tuning will be necessary as more deliveries occur.

### Strengths and Limitations

Strengths of this study include use of modern supervised machine learning techniques in a clinical condition that has not been extensively explored with this approach. This data set represents the largest directly-derived cohort to use these techniques. Additionally, the inclusion of nearly 500 variables in the data set provides a robust cohort from which to create the model, and this size has not been previously seen in the literature. As machine learning methods are centered upon improving performance with increasing inputs, this lends to a superior model. The use of independent validation and test cohorts also supports the strength and lack of bias in our model.

Limitations include the retrospective nature of this study as well as the use of a population from a single tertiary center. Given regional variations in patient populations, our results may not be generalizable to the United States population at large and we do note a higher rate of postpartum hemorrhage in our cohort than previously described. Further validation with an outside cohort is necessary.

The use of the electronic medical record is an additional limitation to our study. Differences/duplications in both location and format of inputs have the potential to impair the accuracy of our abstracted data. The variables related to diagnosis codes are entirely dependent upon provider input and all applicable conditions may not have been entered. However, with a large data set, machine learning algorithms should be able to overcome this deficit as features with a high level of contribution to the outcome should persist when feature selection is implemented.

The class imbalance of positive/negative cases for postpartum hemorrhage in the data set is inevitable given the relatively low incidence of this condition in clinical practice. This was particularly evident in the Support Vector Machine models where every patient was predicted to not be at risk for postpartum hemorrhage. Use of a weighted loss could be considered to compensate for this imbalance.

### Conclusions

In conclusion, machine learning methods are an underutilized approach in obstetrics and can be used to identify women at risk for postpartum hemorrhage who may benefit from individualized preventative measures. Models limited to data available prior to delivery perform nearly as well as those with more complete datasets, identifying nearly three-quarters of patients at risk, supporting their potential utility in the clinical setting.

## Data Availability

The data is available to study collaborators.

## Acknowledgements

The authors wish to thank the members of NYU Langone Health DataCore for their assistance in this project.

